# Improvement of behavioural pattern and alpha-synuclein levels in autism spectrum disorder after consumption of a beta-glucan food supplement in a randomized, parallel-group pilot clinical study

**DOI:** 10.1101/2021.06.28.21259619

**Authors:** Kadalraja Raghavan, Vidyasagar Devaprasad Dedeepiya, Nobunao Ikewaki, Tohru Sonoda, Masaru Iwasaki, Senthilkumar Preethy, Samuel JK Abraham

## Abstract

**Background:** Autism spectrum disorders (ASDs) are a wide range of behavioural disabilities for which there are no definite interventional modalities available. Remedial therapies remain the only option but with varying outcomes. We have evaluated the childhood autism rating scale (CARS) and alpha-synuclein levels in this parallel-group, multiple-arm pilot clinical study after supplementation with a biological response modifier beta-glucan food supplement (Nichi Glucan).

**Methods:** Six subjects with ASD (n = 6) Gr. 1 underwent conventional treatment comprising remedial behavioural therapies and L-carnosine 500 mg per day, and 12 subjects (n = 12) Gr. 2 underwent supplementation with the Nichi Glucan 0.5 g twice daily along with the conventional treatment.

**Results:** There was a significant decrease in the CARS score in all of the children of the Nichi Glucan Gr.2 compared to the control (p-value = 0.034517). Plasma levels of alpha-synuclein were significantly higher in Gr. 2 (Nichi Glucan) than in the control group Gr. 1 (p-value = 0.091701).

**Conclusion:** Improvement of the behavioural pattern CARS score and a correlating alpha-synuclein level, followed by a safe beta-glucan food supplement, warrants further research on other parameters, such as gut-microbiota evaluation, and relevant neuronal biomarkers which is likely to cast light on novel solutions.

## Introduction

Autism spectrum disorders (ASDs) are a group of developmental disabilities that can cause significant impairment in social, emotional, and communication skills (cdc.gov). Several causes and underlying mechanisms have been postulated for the pathogenesis of ASD, including genetic, environmental, immune dysregulation, neuroinflammation, and oxidative stress. Neuronal synaptic imbalance and mutation in synaptic proteins and receptors have also been reported to be associated with ASD (Al-Mazeedi et al., 2020). Synucleins are small soluble proteins that are present in presynaptic terminals, and they regulate synaptic plasticity and neurotransmitter release. Synucleins are important in the context of brains and neurons (Al-Mazeedi et al., 2020; Vargas et al., 2017). Alpha-synuclein has already been reported to be associated with several neurodegenerative disorders, collectively called synucleinopathies such as Alzheimer’s disease (AD), Parkinson’s disease (PD), dementia with Lewy bodies (DLBs), and multiple system atrophy (Al-Mazeedi et al., 2020). Several studies have recently reported a strong association between lower levels of α-synuclein and ASD. At present, there is no definitive cure for ASD (Al-Mazeedi et al., 2020; Kadak et al., 2015; Sriwimol et al., 2018). Interventions involve speech and behavioural therapies to improve the symptoms. According to the research, the microbiota-gut-brain axis is significant because dysbiosis has been observed in gut-related diseases and other generalized disorders, especially of the nervous system, such as AD, multiple sclerosis, PD, and ASD (Srikantha et al., 2019). Therefore, nutritional supplements are considered potential interventions in alleviating gastrointestinal and behavioural symptoms in ASD (Karhu et al., 2020). Beta-glucans, especially yeast-derived ones, have shown a considerable positive outcome as food supplements in modulating gut microbiota (Karhu et al., 2020; Xu et al., 2020). Nichi Glucan is a black yeast–derived AFO-202 beta-glucan that has been in consumption for the past two decades (Ikewaki et al., 2007) and has been shown to have potential as a nutritional supplement to balance metabolic levels of glucose, lipids, and immunomodulators (Dedeepiya et al., 2012; Ganesh et al., 2014; Ikewaki et al., 2021). The present study was undertaken to investigate the effects of Nichi Glucan as a food supplement in children with ASD, especially with relevance to the childhood autism rating scale (CARS) score and alpha-synuclein levels.

## Methods

This study was approved by the institutional ethics committee of the and was registered as a clinical trial in the national clinical trial registry. The caregiver of all the subjects gave their informed consent for inclusion before participation in the study. The study was conducted in accordance with the Declaration of Helsinki.

### Study design

The subjects enrolled in the study had received a diagnosis of ASD by a developmental paediatrician and were verified by a psychologist using a clinical interview for a behavioural pattern that incorporated CARS.

Eighteen subjects (n =18) with ASD in total were enrolled in this prospective, open-label, pilot clinical trial comprising of two arms. The CONSORT flow diagram is presented as Figure 1.

**Figure 1:**
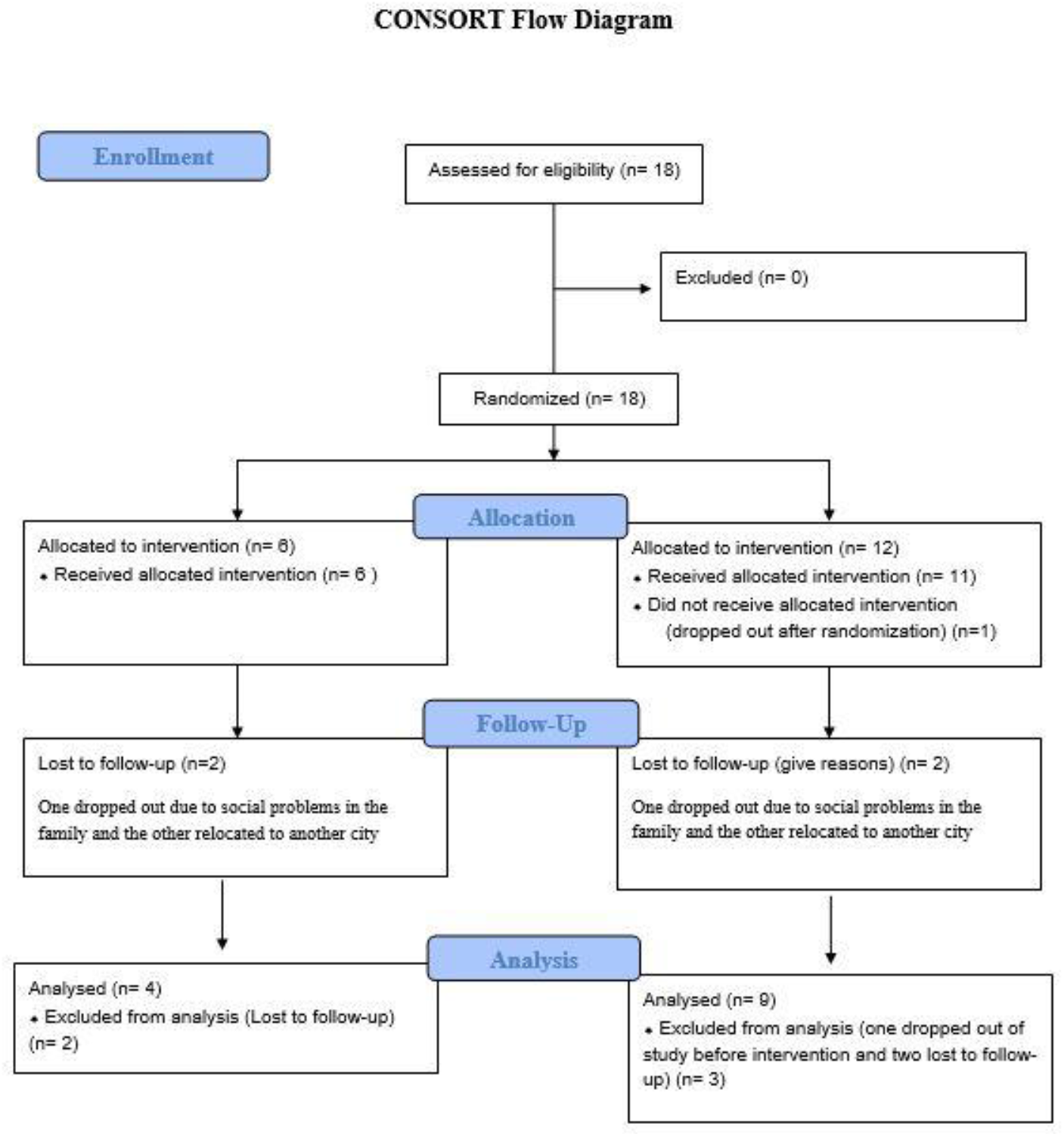
CONSORT Flow diagram for description of the clinical trial.

Arm 1 or Gr. 1: Control: Six subjects with ASD (n = 6) underwent conventional treatment comprising remedial behavioural therapies and L-Carnosine 500 mg per day.

Arm 2 or Gr. 2: Treatment arm: 12 subjects (n = 12) underwent supplementation with Nichi Glucan food supplement along with conventional treatment. Each subject consumed two sachets (0.5 g each) of Nichi Glucan daily—one sachet with a meal twice daily—for a period of 90 days.

### Inclusion criteria

i. Age: 3 to 18 years
ii. Gender: Both male and female
iii. ASD criteria as per CARS score
iv. Parents/caretakers willing to provide consent for their children to actively participate in the study

### Exclusion criteria

i. Subjects aged more than 18 years old
ii. Any child with acute general illness or who has been on any antibiotic, anti-inflammatory, or antioxidant treatment in the two weeks prior to enrolment in the study
iii. Hyperallergic to any of the investigational products
iv. Subjects with long-standing infections

### Outcome measures

#### i. Childhood autism rating scale (CARS)

The CARS was monitored at baseline and after 90 days between the Gr.1 (control) and Gr.2 (Nichi Glucan). The CARS score was calculated based on a cumulative score obtained on the CARS scale, wherein a score below 30 indicates absence of sufficient signs and symptoms indicative of autism, a score between 30 and 36 indicates mild-to-moderately severe autism, and a score from 37 to 60 is correlated with severe autism (Ramaekers et al., 2019). The psychologist who performed the assessment and the parents were blind to the participant’s treatment status; hence, this is a double-blind study.

#### ii. Evaluation of plasma alpha-synuclein

Human alpha-synuclein (α-syn) levels in plasma were measured in peripheral blood at baseline and after the study’s completion at 90 days. The measurement was performed using the human α-synuclein (α-syn) ELISA kit (KINESISDx, USA) as per the manufacturer’s instructions.

### Data analysis

All data were analysed using Excel software statistics package analysis software (Microsoft Office Excel®); Student’s paired t-tests were also calculated using this package; and p-values < 0.05 were considered significant.

## Results

During enrolment, six subjects with ASD (n = 6) could be enrolled in the control group (Gr. 1), whereas in the treatment group (Gr. 2), one of them dropped out before the start of the study. During the study, four subjects were lost to follow-up: two in Gr. 1 (one dropped out due to social problems in the family, and the other relocated to another city) and two in Gr. 2 (one dropped out due to social problems in the family, and the other relocated to another city). A total of 13 subjects (four in Gr. 1 and nine in Gr. 2) completed the study. One female subject was in both Gr. 1 and Gr. 2. The rest were male.

### Adverse effects

Only one child exhibited possible mild adverse effects related to increased bowel movements in Gr. 2 for one week after supplementation with Nichi Glucan, which settled on its own. No adverse effects were found in any of the other children.

### Score on CARS scale

Among the children in the control group (Gr.1), all four were in the category of severe autism, and their score at baseline ranged from 37 to 52 (mean = 42.75 ± 5.76). Among the nine children in Gr.2, two were in the mild-to-moderate category of autism (mean = 33.5 ± 2.5), whereas the remaining seven were in the category of severe autism (mean = 43.71 ± 4.80). After the intervention, the mean CARS score in the four children of the control group was 42.5 ± 5.4, while in Gr.2 (Nichi Glucan), the mean of the CARS score in the two children with mild-to-moderate autism was 32.5 ±0.5. In the remaining seven children, the CARS score after Nichi Glucan intervention had a mean of 40.1 ± 5.96. Thus, there was a significant decrease in the CARS score in all of the children in the Nichi Glucan Gr.2 group compared to the control (p-value = 0.034517), with an average of 3 points in the improvement of autism’s signs and symptoms, whereas the improvement was very mild or nil in Gr.1 (Figure 2). Among the various parameters assessed on the CARS, there was visible subjective improvement in the emotional response, including reduction in irritability and anger (88%), sleep improvement (88%), speech characteristics with improvement in finger pointing and monosyllables in 77%, and improved responses to the caregiver in 77% of the children in Nichi Glucan Gr. 2, but these improvements were very mild or nil in Gr.1.

**Figure 2:**
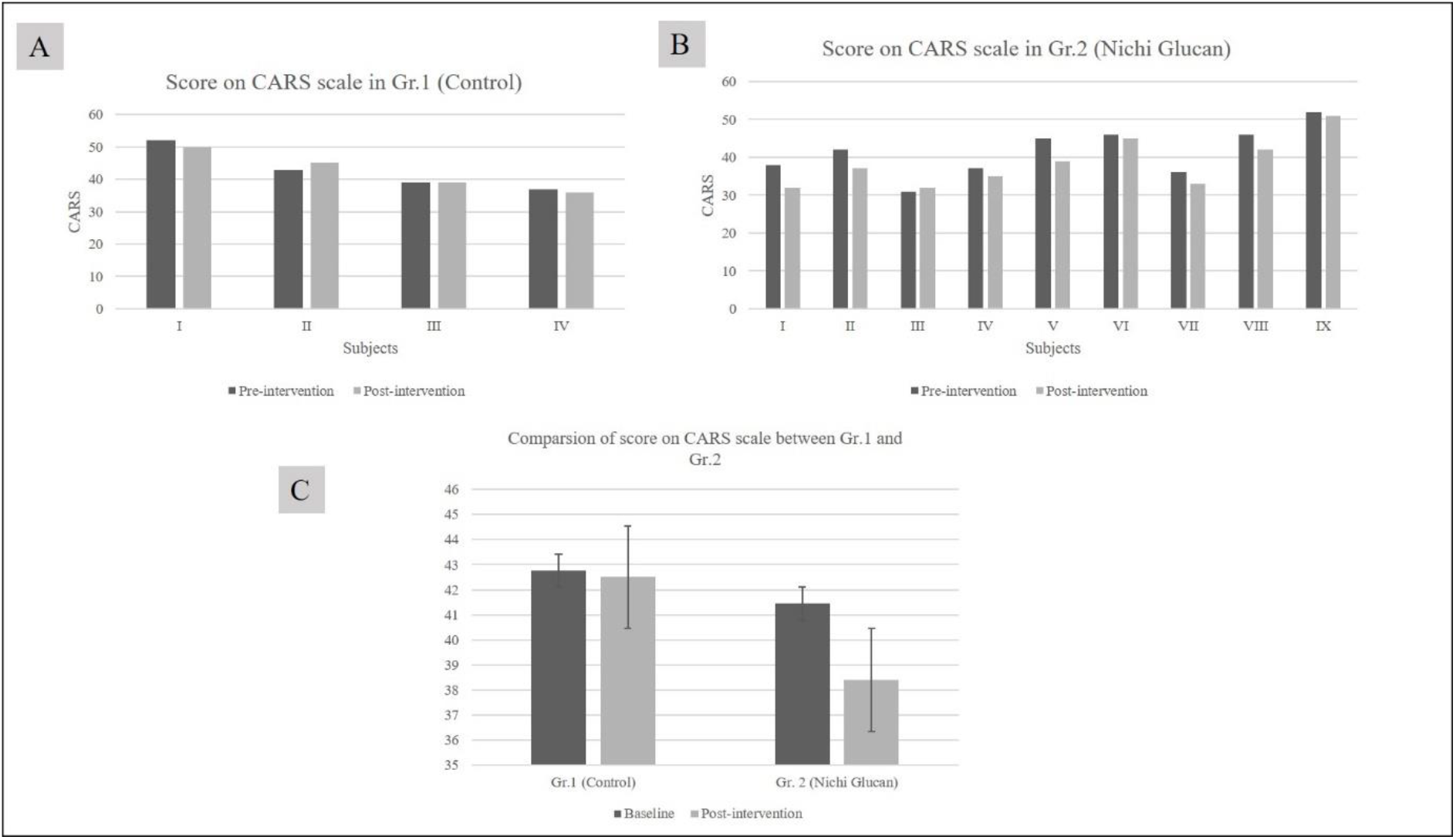
A. The score on the childhood autism rating scale (CARS) in Gr.1 (control), which was very mild and showed no improvement after the study, whereas the score decreased in all children by an average of 3 points for B. Gr.2 (Nichi Glucan). C. Comparison between Gr.1 and Gr.2 showed a significant decrease in the CARS score, indicating improvement in autism’s signs and symptoms in Gr.2 (Nichi Glucan) compared to Gr.1 (control) post-intervention (p-value = 0.034517).

Plasma levels of alpha-synuclein ranged between 0.12 and 20.41 ng/dl (mean = 9.73 ng/dl) in the control group and between 0.45 and 41.12 ng/dl (mean = 9.39 ng/dl) in the treatment group at baseline. After the intervention, plasma levels of alpha-synuclein increased, with a mean increase in levels of 26.72 ng/dl in the treatment (Nichi Glucan) Gr.2 group compared to the control group Gr. 1 (mean increase = 10.56 ng/dl) (p-value = 0.091701) (Figure 3).

**Figure 3:**
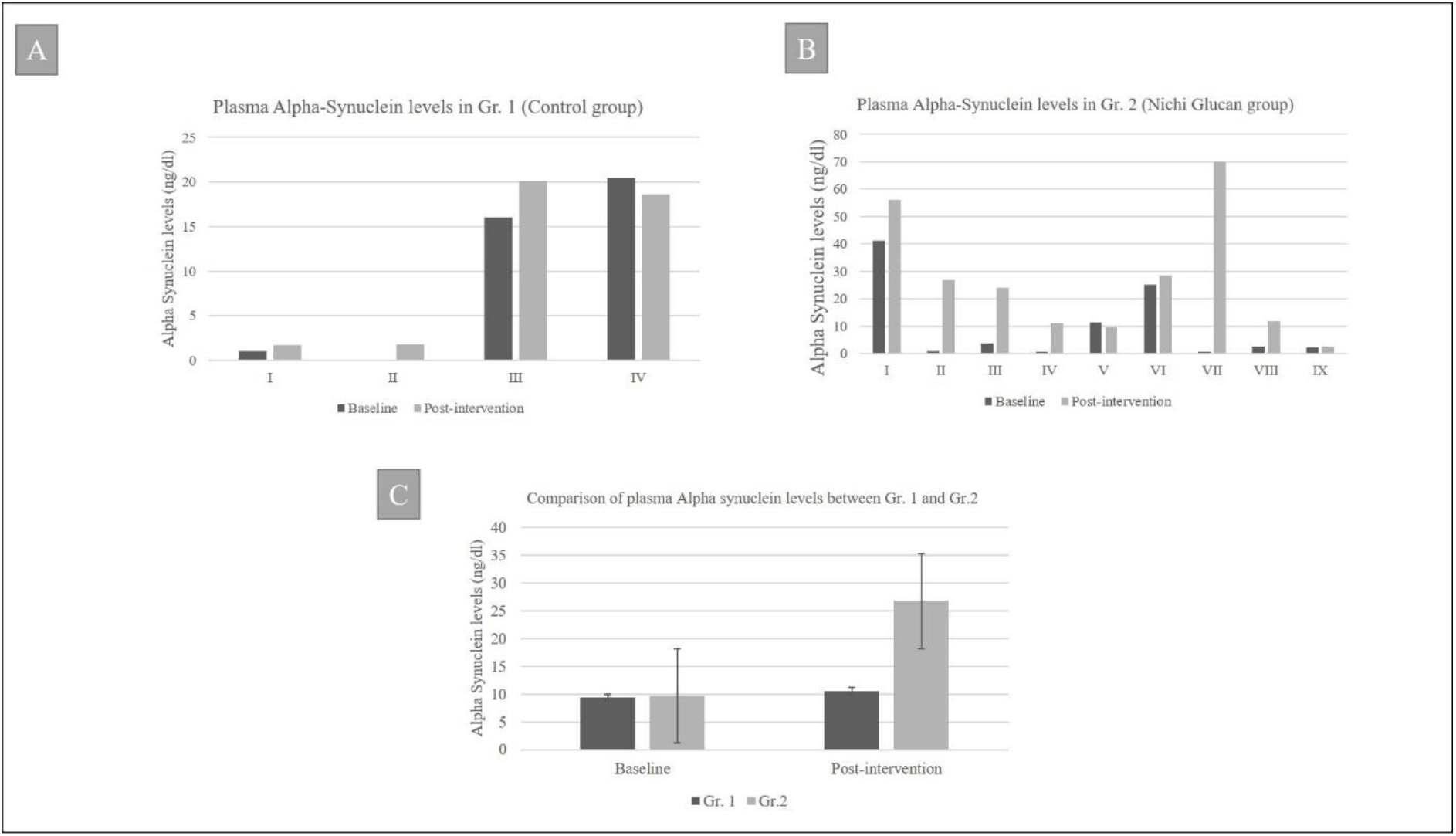
A. Plasma alpha-synuclein levels in Gr. 1 (control) pre- and post-intervention. B. Plasma alpha-synuclein in Gr. 2 (Nichi Glucan) pre- and post-intervention with C. Plasma alpha-synuclein levels in Nichi Glucan group Gr. 2 showed a significant increase compared to Gr.1 (p-value = 0.091701).

## Discussion

In this study of 13 subjects, the behavioural pattern evaluated by the CARS score improved in all nine subjects of Gr.2 (Nichi Glucan) (Figure 1), especially on the emotional aspects and sleep-related parameters, and the alpha-synuclein levels increased significantly in these nine subjects compared to the control (Figure 2). Alpha-synuclein plays a key role in the synaptic functions of neurons by regulating CADPS2 mRNA expression. There are reports that the neural overconnectivity and synapse alteration associated with the pathogenesis of ASD may actually owe their aetiology to alpha-synuclein dysregulation (Kadak et al., 2015; Sriwimol et al., 2018; Obergasteiger et al., 2014). Further, alpha-synuclein has recently been considered one of the important biomarkers for the diagnosis of autism and ASD, wherein the levels are low compared to age-matched controls (Kadak et al., 2015; Sriwimol et al., 2018; Siddique et al. 2020). In regard to neurodegenerative diseases such as PD, the reports have been varied, with some reporting lower than normal levels and others higher. In a correlating hypothesis of the plasma alpha-synuclein level, between autism and neurodegenerative diseases, it has been proposed that alpha-synuclein aggregation in the neural synapse may lead to lower plasma levels (Sriwimol et al., 2018). Whether the increase in alpha-synuclein levels in plasma in the ASD patients after Nichi Glucan supplementation is due to regulation/prevention of alpha-synuclein’s aggregation in the neural synapse must be investigated because an earlier study on beta-glucan from yeast showed reduction in alpha-synuclein expression on the brain substantia nigra in Parkinson’s rat model (Masruroh et al., 2017). However, no single mechanism, intervention, or therapy has proven its ability to regulate alpha-synuclein levels, especially in children with ASD. In our study, which is the first of its kind, the plasma alpha-synuclein levels showed significant increase after Nichi Glucan supplementation, and the levels were in line with those that were reported for children without ASD (Kadak et al., 2015; Sriwimol et al., 2018).

Studies on children with ASD have indicated there is an underlying neuroinflammatory process occurring in different regions of the brain involved in microglial activation, thus resulting in a loss of connections or underconnectivity of neurons and leading to behavioural manifestations (Shah et al., 2009). MCP-1, IL-6, IL-10, and TNF-α have been shown to be expressed in higher levels in children with autism (Shah et al., 2009). Beta-glucan has been proven to reduce the expression of inflammatory and proinflammatory markers, including Il-6 and TNF-α (Ikewaki et al., 2007), apart from having a neuroprotective effect by attenuating inflammatory cytokine production through microglia (Alp et al., 2012). This mechanism of counteracting ASD inflammation by Nichi Glucan supplementation deserves further research.

In another study, beta-glucan reduced induced microglia activation and its phagocytosis of synaptic puncta and upregulation of proinflammatory cytokine (TNF-α, IL-1β, and IL-6) mRNA expression apart from promoting Tau signalling and improving cognition and brain function via the gut-brain axis (Shi et al., 2020). The mechanism by which the beta-glucan promoted behavioural improvement in the present study and correlated with the regulation of alpha-synuclein levels needs further in-depth research, not only for ASD but also for neurodegenerative diseases such as AD, PD, and so on, especially with regard to its effects on the gut-microbial ecosystem. The evolving data on the gut-brain axis and gut microbiota indicate there are two major approaches to balancing gut microbiota: probiotic and prebiotic. Probiotic approaches, such as nutritional probiotics, faecal transplantation, and so on, involve direct administration of the beneficial microorganisms that have to colonise the gut (Peng et al., 2020). However, the gut environment must be conducive for such probiotic supplementation. This is where prebiotic approaches come in, such as Nichi Glucan, which help in regulating the gut-microbial ecosystem and preventing chronic inflammatory status Peng et al., 2020); this must be validated by future studies in terms of the effects of Nichi Glucan and gut microbiota in their relevance to ASD.

The limitation of the study is the limited number of participants, the unequal distribution of genders, and the number of participants between the groups. However, this is only a pilot study, and larger randomized, multi-centric clinical trials are warranted. Nevertheless, the study is significant as it has identified a simple nutritional supplemental intervention based on a naturally derived beta-glucan, the Nichi Glucan, which could stimulate endogenous alpha-synuclein secretion, promote better synaptic regulation, and improve the behaviour symptoms of children with autism. However, the results suggest that the benefits will be considerable when evaluated in terms of social and emotional well-being and alleviation of caregiver stress, which is extremely significant.

## Conclusion

Patients with ASD showed improvement in behavioural symptoms and improved levels of plasma alpha-synuclein; thus, this pilot clinical study of nutritional supplementation with an AFO-202 strain of black yeast *aureobasidium pullulans* produced the biological response modifier beta-glucan (Nichi Glucan). Evaluation as per the CARS score has also shown significant beneficial effects. Although further validations need to be performed, the study definitely confirms the potential of Nichi Glucan as a simple but effective food supplement to be considered as a routine in children with ASD. Further research on the mechanisms of its action in improving alpha-synuclein levels and balancing the immune system in the context of managing chronic inflammation and gut-microbiota regulation as a prebiotic is likely to improve understanding of other diseases caused by neuroinflammation such as PD and AD.

## Data Availability

The authors confirm that the data supporting the findings of this study are available within the article

## Acknowledgements

The authors thank

1. Mr. Mohan Ponnusamy & staff of Kenmax for their assistance during the clinical study and data collection of the manuscript.
2. Mr. Takashi Onaka, Mr. Yasunori Ikeue, Mr. Mitsuru Nagataki (Sophy Inc, Kochi, Japan), for necessary technical clarifications.
3. Loyola-ICAM College of Engineering and Technology (LICET) for their support to our research work.

## Notes

### Competing Interest Statement

Author Samuel Abraham is a shareholder in GN Corporation, Japan which in turn is a shareholder in the manufacturing company of the AFO 202 Beta Glucan.

### Clinical Trial

CTRI/2020/10/028322

### Funding Statement

No external funding was received for the study

### Author Declarations

The study was approved by the Institutional Ethics Committee (IEC) of Saravana Multispeciality Hospital, Madurai, India on 24th August, 2019.

